# Artificial Intelligence Detects Cardiac Amyloidosis Before Clinical Diagnosis

**DOI:** 10.64898/2026.03.03.26347432

**Authors:** Victoria Yuan, Saisanjana Kalagara, Hirotaka Ieki, Ankeet S. Bhatt, Andrew P. Ambrosy, Aly Elezaby, Lily K. Stern, Paul P Cheng, David Ouyang

**Author notes:** **Corresponding Author:** David Ouyang, 4480 Hacienda Drive, Pleasanton, CA 94588.

## Abstract

The approval of novel disease-modifying treatments for cardiac amyloidosis (CA) offers an avenue to stabilize disease progression. Timely diagnosis of CA is imperative so that patients can be connected to treatment earlier in their course. Echocardiography is a widely available, non-invasive modality to screen for CA. However, diagnosis of CA is often delayed due to phenotypic mimickers. Artificial intelligence (AI) applied to echocardiography may facilitate early detection and connect patients to more definitive diagnostic testing. In this case report, we analyzed the frequency of echocardiography prior to clinical diagnosis of amyloidosis and deployed an externally validated AI model in 349 patients with documented amyloidosis at Cedars-Sinai Medical Center. On average, AI detection was positive in patients with documented CA 218 days prior to clinical diagnosis and 209 days in all-comers. When integrated with clinical histories, AI may facilitate earlier detection of CA and connect patients to more timely management.

## Introduction

Cardiac amyloidosis (CA) causes contractile dysfunction through gradual deposition of amyloid fibrils within myocardium. However, diagnosis is often delayed given phenotypic mimickers^1^. The approval of novel disease-modifying treatments offers an avenue to stabilize disease progression, with greater clinical impact earlier in the course^2,3^. Timely detection is imperative to connect patients to disease-specific management. Echocardiography is the most common form of cardiac imaging. Artificial intelligence (AI) applied to echocardiography may improve precision diagnostics of CA by differentiating it from mimickers^4^. In this report, we evaluated the frequency of echocardiography prior to clinical diagnosis and investigated whether a validated AI model can facilitate early detection of CA^4^.

## Methods

Using ICD 9/10 codes and manual chart review, we identified patients with a clinical diagnosis of amyloidosis, with and without cardiac involvement, who received care at Cedars-Sinai Medical Hospital (CSMC) between 2006-2024. We evaluated the incidence of echocardiography imaging prior to clinical diagnosis. Echocardiography studies prior to clinical diagnosis were evaluated with an externally validated, open-source AI model that predicts the probability of cardiac amyloidosis^4^. There was no overlap between the patients in the model development cohort at Stanford Healthcare and our external validation cohort at CSMC. This study was approved by the institutional review board at CSMC.

## Results

A total of 396 patients (99 [25.0%] female; age 72.2 [SD 10.4] years) were identified to have a clinical diagnosis of amyloidosis at CSMC during this time, of which 304 (76.8%) patients (80 [26.3%] female; mean [SD] age 73.8 [10.2] years) had echocardiography studies prior to initial diagnosis. The median time between echocardiography and initial diagnosis was 131 days. 186 (61.2%) patients had AL amyloidosis, 95 (31.3%) had transthyretin amyloidosis (ATTR), 10 (3.3%) had non-cardiac organ-limited amyloidosis, 6 (2.0%) had AA amyloidosis, 6 (2.0%) had dialysis-related amyloidosis, and 1 (0.3%) had ALECT2 amyloid. Of these 304 patients, 221 (72.7%) (43 [19.5%] female; mean [SD] age 75.3 [10.0]) were recognized to have cardiac involvement, with 124 (56.1%) having AL amyloidosis, 94 (42.5%) with ATTR, and 3 (1.4%) with dialysis-related amyloidosis. Patients with echocardiography before diagnosis were more likely to be older (one-sided t-test p-value < 0.001) than those without. There was no difference in the proportion of Black (p = 0.19), Asian (p = 0.37), or Hispanic (p = 0.65) patients. Of the 304 all-comers, 233 (76.6%) patients had at least 2 serial echocardiography studies; 175 (79.2%) of the 221 patients with CA had at least 2 studies.

Only 7.7% of the prior studies had possible amyloidosis listed as an indication for imaging, with the most common indications for heart failure (20.2%), valvular disease (17.7%), atrial fibrillation (11.3%), dyspnea (10.8%), chest pain (9.2%), pericardial disease (8.7%), cardiomyopathy (8.4%), coronary artery disease (6.9%), pre-transplant evaluation (5.5%), and other (6.8%). Patients diagnosed with CA had similar indexed end-systolic volume (19.5 vs 19.7 mL/m^2^, p = 0.90) and indexed end-diastolic volume (42.9 vs 39.7, p = 0.19), but decreased LVEF (57.3% vs 54.1%, p = 0.01), increased IVSd (1.13 vs. 1.52 cm, p < 0.001) and increased LVPWd (1.12 vs. 1.44 cm, p < 0.001) compared to patients without documented cardiac involvement.

AI detection scores were positive in 90 (40.7%) CA patients prior to clinical diagnosis and positive in 100 (32.9%) all-comer amyloidosis patients (**Figure 1**). Of the all-comers with early AI detection, 61 (61.0%) patients had AL amyloidosis, 35 (35.0%) had ATTR, 3 (3%) had dialysis-related amyloid, and 1 (1.0%) had AA amyloidosis. Within the CA cohort with early AI detection, 53 (58.9%) had AL amyloidosis, 34 (37.8%) had ATTR, and 3 (3.3%) had dialysis-related amyloidosis. In the cohort of patients with clinically diagnosed CA, 22 (31.4 %) patients were identified by AI at least 1 year prior to clinical diagnosis and 30 (31.9%) 6 months prior from imaging alone. In all-comers, 25 (22.3%) patients were identified 1 year prior to clinical diagnosis and 34 (23.3%) 6 months prior. Overall, AI prediction was positive with a mean of 273 days before clinical diagnosis of CA and 260 days before diagnosis in all-comers (**Figure 1**). Echocardiography 3 to 6 months prior to clinical diagnosis demarcated an inflection point in model sensitivity, with the proportion of positive cases similar to that within one month of diagnosis.

**Figure 1.**
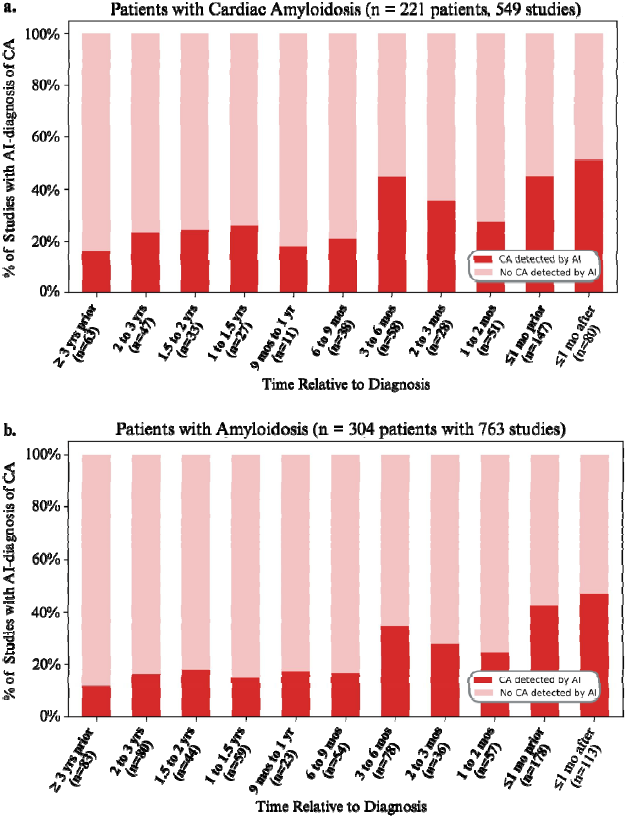
Percent of echocardiography studies that were detected as positive cases by AI-cardiac amyloidosis scores based on time relative to clinical diagnosis in patients diagnosed with **a)** cardiac amyloidosis and **b)** amyloidosis; time intervals prior to diagnosis are bolded.

## Discussion

Patients often have echocardiography performed prior to clinical diagnosis of CA, with the initial study often having signs of CA. Our analysis demonstrated that before clinical diagnosis, echocardiography was performed more commonly to evaluate patients for heart failure or valvular disease than amyloidosis. Using AI re-review of historical echocardiography, AI frequently suspected CA six months to one year before clinical diagnosis. Model performance was even higher in patients diagnosed with cardiac involvement. When integrated with patient histories, AI can potentially facilitate earlier identification of CA, which can in turn connect patients earlier to disease-specific treatment and improve their long-term prognoses.

## Data Availability

Due to its potentially identifiable nature, data in the present study will not be made available.

## Abbreviations

1. CA: cardiac amyloidosis
2. AI: artificial intelligence
3. A4C: apical 4-chamber
4. AUC: area under the receiver operator characteristic curve
5. ATTR: transthyretin amyloidosis
6. LV: left ventricular
7. IVSd: diastolic interventricular septal wall thickness
8. LVPWd: diastolic left ventricular posterior wall thickness

## Notes

**Disclosures:** Dr. Ambrosy reports research support through grants from the National Institutes of Health (R01HL173866 and R01AG091005), the Food and Drug Administration (U01FD008704), The American Heart Association, The Permanente Medical Group, Kaiser Permanente Northern California Community Benefits Programs, Garfield Memorial Fund, Abbott Laboratories, Abiomed, Amarin Pharma, Inc., Bayer Pharma AG, Boehringer Ingelheim, Cordio Medical, Edwards Lifesciences LLC, Esperion Therapeutics, Inc., Merck, and Novartis. Consulting for Bristol Meyer Squibb, Bayer Pharma AG, Corstasis, Merck, Novo Nordisk, Pharmacosmos A/S, Pfizer, scPharma, SQ Innovation, Reprieve Cardiovascular, and VisCardia. Ankeet S. Bhatt has received research grant support to his institution from National Institutes of Health/National Heart, Lung, and Blood Institute, National Institutes of Health/National Institute on Aging, American College of Cardiology Foundation, American Heart Association, Sanofi Pasteur, Abiomed, AstraZeneca and the Centers for Disease Control and Prevention and consulting fees or equity from Heart Health Leaders, Sanofi Pasteur, Merck, Amgen, AstraZeneca, Esperion, Bayer, Cytokinetics, and Novo Nordisk and Eko Health. Dr. Stern has received research support from Intellia Therapeutics and Pfizer; advisor fees from Eidos/BridgeBio and Pfizer; honoraria fees from Alnylam Pharmaceuticals. Dr. Ouyang reports consulting, equity, or honoraria for lectures from AstraZeneca Alexion, EchoIQ, Ultromics, Pfizer, InVision, the Korean Society of Echocardiography, and the Japanese Society of Echocardiography. The remaining authors have nothing to disclose.

**Funding:** Ms. Yuan gratefully acknowledges funding from the Sarnoff Cardiovascular Research Foundation. Dr. Ouyang reports support from the National Institute of Health (NIH; NHLBI R00HL157421, R01HL173487 and R01HL173526), AstraZeneca, and Alexion.

### Competing Interest Statement

Dr. Ambrosy reports research support through grants from the National Institutes of Health (R01HL173866 and R01AG091005), the Food and Drug Administration (U01FD008704), The American Heart Association, The Permanente Medical Group, Kaiser Permanente Northern California Community Benefits Programs, Garfield Memorial Fund, Abbott Laboratories, Abiomed, Amarin Pharma, Inc., Bayer Pharma AG, Boehringer Ingelheim, Cordio Medical, Edwards Lifesciences LLC, Esperion Therapeutics, Inc., Merck, and Novartis. Consulting for Bristol Meyer Squibb, Bayer Pharma AG, Corstasis, Merck, Novo Nordisk, Pharmacosmos A/S, Pfizer, scPharma, SQ Innovation, Reprieve Cardiovascular, and VisCardia.
Ankeet S. Bhatt has received research grant support to his institution from National Institutes of Health/National Heart, Lung, and Blood Institute, National Institutes of Health/National Institute on Aging, American College of Cardiology Foundation, American Heart Association, Sanofi Pasteur, Abiomed, AstraZeneca and the Centers for Disease Control and Prevention and consulting fees or equity from Heart Health Leaders, Sanofi Pasteur, Merck, Amgen, AstraZeneca, Esperion, Bayer, Cytokinetics, and Novo Nordisk and Eko Health.
Dr. Stern has received research support from Intellia Therapeutics and Pfizer; advisor fees from Eidos/BridgeBio and Pfizer; honoraria fees from Alnylam Pharmaceuticals.
Dr. Ouyang reports consulting, equity, or honoraria for lectures from AstraZeneca Alexion, EchoIQ, Ultromics, Pfizer, InVision, the Korean Society of Echocardiography, and the Japanese Society of Echocardiography. The remaining authors have nothing to disclose.

### Funding Statement

Ms. Yuan gratefully acknowledges funding from the Sarnoff Cardiovascular Research Foundation. Dr. Ouyang reports support from the National Institute of Health (NIH; NHLBI R00HL157421, R01HL173487 and R01HL173526), AstraZeneca, and Alexion.

### Author Declarations

This study was approved by the Institutional Review Board at Cedars-Sinai Medical Center.

